# MATRIX METALLOPROTEINASE 26 (MMP-26) OVEREXPRESSION IN PROSTATIC ADENOCARCINOMA

**DOI:** 10.1101/2021.05.28.21257789

**Authors:** Romildo Luciano da Silva, Ingrid Tavares de Lima, Francisco Luís Almeida Paes, Sandra Maria Souza da Silva, Ana Pavla Almeida Diniz Gurgel, Fabiano Santos, Jacinto da Costa Silva Neto

**Affiliations:** Pathological Anatomy Laboratory, Clinical Hospital of Federal University of Pernambuco (UFPE), Brazil; Cytological and Molecular Research Laboratory, Federal University of Pernambuco (UFPE), Brazil; Department of Engineering and Environment, Federal University of Paraiba, Paraiba, Brazil; Technology and Innovation Program, International Development Research Centre (IDRC), Ottawa, Canada; Department of Histology and Embryology (UFPE), Recife, Pernambuco, Brazil

**Keywords:** Matrix Metalloproteinase-26 (MMP-26), Immunohistochemistry (IHC), Prostate-Specific Antigen (PSA), Prostate cancer

## Abstract

**Introduction:** Matrix metalloproteinases (MMP) have been identified as biomarkers for several diseases, including cancer. MMP-26 is constitutively expressed in some cancer cells of epithelial origin. Despite this, there is a lack of studies regarding the expression of MMP-26 on prostatic carcinoma.

**Aim:** Here, we investigate the expression of the MMP-26 peptide in benign and malign prostatic tissues.

**Patients and Methods:** For this, 150 specimens, including atrophy (N = 25), prostatic intraepithelial neoplasia (PIN) (N = 25), benign prostatic hyperplasia (BPH) (N = 50), and prostatic adenocarcinoma (PA) (N = 50), were immunohistochemically (IHC) examined for the expression of MMP-26.

**Results:** MMP-26 expression was positive in 70 (46.7%) out of the 150 samples, being more prevalent in the PA group (46/50 cases,92%), followed by PIN (22/25 cases, 88%). The BPH group showed only 2/50 (4%) positive cases, and the atrophy group showed no reactivity. ROC curve analysis showed that MMP-26 immunoexpression had a higher area under the curve between PA vs atrophy+PIN+BPH (AUC=0.94; 95% CI 0.9-0.98), PA+PIN vs atrophy+BPH (AUC=0.97; 95% CI 0.94-0.99) and PA vs atrophy+BPH (AUC=0.97; 95% CI 0.95-1.00) groups. In addition, the expression and intensity of the MMP-26 reaction showed a significant association with total PSA values (*P*=0.001).

**Conclusions:** Our results showed that MMP-26 immunoexpression was useful to differentiate a group of benign and malignant samples in prostate tumors. This characteristic could assist in the predictive assessment and, consequently, in the development of new strategies for the diagnosis, prognosis, and treatment of prostate cancer.

## INTRODUCTION

Prostate cancer (PC) is one of the most prevalent cancers worldwide, ranking second among malignant neoplasms that affect men ^1^. PC screening tools include serum prostate-specific antigen (PSA) measurement, rectal examination, and biopsy. Despite its name, PSA has low specificity presenting with high levels in benign conditions such as prostatitis and hyperplasia, which results in unnecessary treatment and elevated rates of negative biopsy results ^2^. Therefore, research of new biomarkers for early, differential diagnosis and with potential for PC prognostic value has been undertaken.

Matrix metalloproteinase (MMPs) are endopeptidases that promote degradation and remodeling of the extracellular matrix and play an essential role in various physiological and pathological processes ^3^. Its exacerbated expression is associated with invasion, angiogenesis, metastasis, and poor prognosis of several types of cancer, such as thyroid ^4^, breast ^5^, and prostate ^6^. MMP-26 is the smallest molecule in the MMP family that was recently discovered. Its constitutive expression was described in the endometrium and placenta, as well as in cancer cells of epithelial origin ^7^. Its primary substrates include fibronectin, fibrinogen, vitronectin, laminin, and type IV collagen. MMP-26 also can activate pro-MMP-9, known to promote invasion and angiogenesis in several human tumors ^8^.

High expression of MMP-26 has been demonstrated in breast, lung, liver, endometrial, ovarian, and esophageal cancer cell lines ^9–14^. Nevertheless, research on its role in PC is still scarce. Recently, the prognostic value of MMP-26 has been evaluated in colorectal cancer, where significant expression was associated with invasion, metastasis, and lower overall patient survival ^15^. MMP-26 was also significantly higher in both serum and tissue of patients with PC ^16^. Despite this, there are fewer studies devoted to the MMP-26 and PC. Hence, we investigate the expression of the MMP-26 in benign and malign prostatic tissues and its relation with PSA as putative biomarkers for differentiating PC patients from atrophy, prostatic intraepithelial neoplasia (PIN), and benign prostatic hyperplasia (BPH) patients.

## PATIENTS AND METHODS

### Patients

One hundred and fifty histological samples were obtained from patients presenting with prostatic lesions and treated at the *Clinical Hospital of Federal University of Pernambuco* (HC/UFPE). The samples were divided into 4 groups, based on its histopathological classification: prostatic atrophy (N = 25), prostatic intraepithelial neoplasia (PIN) (N = 25), benign prostatic hyperplasia (BHP) (N = 50) and prostatic adenocarcinoma (PA) (N = 50). Tumor samples in paraffin blocks were obtained from the Pathology department of the hospital along with their respective clinical reports. Clinical data such as PSA dosage, patient age, and ethnicity were obtained from medical records. The UFPE research ethics board reviewed the protocol and approved the study (approval number CAAE 79701517100005208).

### Immunohistochemistry analysis

Blocks containing representative parts of the tumor were subjected to microtomy generating 5µ slices, followed by processing and reactivity. Each slide was deparaffinized in xylol, bathed in decreasing solutions of ethyl alcohol (100%, 100%, and 70%), and deionized water for 5 minutes. After antigenic recovery with citrate buffer (10 mM, pH 6.0) and blocking of endogenous peroxidase and other proteins using the Envision Flex kit (Dako Denmark A / S), samples were incubated with anti-MMP-26 (FNab 05242, Fine Test, 1: 500) in a humid chamber, overnight at 4°C. The material was subsequently immersed in phosphate-buffered saline (PBS, 100 mM, pH 7.2) and incubated with the HRP polymer (Horseradish peroxidase). After PBS wash, the samples were incubated with the Diaminobenzidine developer (DAB), counterstained with hematoxylin, immersed in running water, and increasing alcoholic solution (70%, 100%, and 100%) for 5 minutes each. Finally, the slides were immersed in xylol and assembled with coverslips interfaced with synthetic resin. Negative controls were performed using the same protocol in the absence of the primary antibody. Endometrial adenocarcinoma samples were used for positive controls.

### Image Collection and Analysis

Labeled cells were observed using a Zeiss AxioCam microscope and a Zen Blue image capture system (edition 2011). Manual quantification was used where the parenchymal epithelial cells were evaluated. The reaction was considered as negative when less than four epithelial cells were marked per field, and positive when more than four marked cells were observed per field. The intensity of the reaction was evaluated semiquantitatively according to the reaction in the nucleus or cytoplasm of the cells: weak (04 to 09 nuclei or reactive cytoplasms), moderate (10 to 15 nuclei or reactive cytoplasms), and intense (above 15 nuclei or reactive cytoplasms).

### Statistical Analysis

Categorical variables were analyzed by the Chi-Square test, the Fisher’s Exact Test, and the Likelihood Ratio Test, according to data distribution. Numerical variables were analyzed with the Kruskal-Wallis test. Positive and negative IHC were compared through the Mann-Whitney test. Area under a receiver operating characteristic (ROC) was calculated to estimated sensitivity and specificity for tests. Statistical significance was judged at the 0.05 level. IMB-SPSS version 23 was used to perform the analyses.

## RESULTS

### Characteristics of the study population

The age of the study population ranged from 41 to 88 years, with a mean of 66.8 years of age. Patients aged 60 to 69 years represented the most prevalent age group (45.3%), followed by patients aged 70 or over (36.7%), with the remaining 18% having between 41 to 59 years of age (*P*=0.447). Ethnicity was different between groups, being patients identified as mixed-race comprised the most prevalent ethnic group (44%), followed by 19.3% of afro-Brazilians and Caucasian (10%) (*P*=0.010). Ethnic information was absent for 26.7% of the sample. Regarding risk habits for prostatic diseases, 29.3% of the participants reported alcohol consumption (*P*=0.286) and 32.7% were smokers (*P*=0.649).

### Analysis of patients’ PSA levels

Measurements of the patient’s PSA levels were obtained from medical records and divided according to serum levels: up to 4 ng/ml (24.7%), 4 to 10 ng/ml (48%), greater than 10 ng/ml (18%). PSA levels were absent for 14 patients. PA samples presented with the highest percentages of PSA levels (26.7%), followed by BHP (19.3%), and atrophy (17.3%) (Table 2). Table 3 summarizes PSA levels in the study population. A significant difference was observed between groups (*P* =0.002). The percentage of PSA value in the range of 4 to 10 was higher in the group with atrophy (69.6%) and lower in the BHP group (40.0%), while the percentage with PSA up to 4 was higher in the BHP group (47.5%), lower in the PA group (12.5%) and varied from 24.0% to 26.1% in the remaining two groups. The mean and median of total PSA were correspondingly higher in the PA group when compared to the other groups, with the lower occurring in the atrophy group. We observed a significant difference between the PA group and the other remaining groups.

**Table 1:** Characteristics of the study population according to histopathological group

| Variables (n= 150) | Group |  |  |  |  |  |  |  | p-value |
| --- | --- | --- | --- | --- | --- | --- | --- | --- | --- |
|  | Atrophy (n=25) |  | PIN (n=25) |  | BPH (n=50) |  | PA (n=50) |  |  |
|  | n | % | n | % | n | % | n | % |  |
| Age group (years) |  |  |  |  |  |  |  |  | p<sup>(1)</sup> = 0.447 |
| 41 a 59 (n=27) | 3 | 12 | 5 | 20 | 12 | 24 | 7 | 14 |  |
| 60 a 69 (n= 68) | 12 | 48 | 11 | 44 | 17 | 34 | 28 | 56 |  |
| 70+ (n=55) | 10 | 40 | 9 | 36 | 21 | 42 | 15 | 30 |  |
| TOTAL | 25 | 100 | 25 | 100 | 50 | 100 | 50 | 100 |  |
| Ethnicity/color |  |  |  |  |  |  |  |  | p<sup>(2)</sup> = 0.010* |
| White (n=15) | 1 | 4,8 | 3 | 17.6 | 3 | 7.0 | 8 | 27.6 |  |
| Brown (n= 66) | 9 | 42.9 | 9 | 52.9 | 30 | 69.8 | 18 | 62.1 |  |
| Black (n = 29) | 11 | 52.4 | 5 | 29.4 | 10 | 23.3 | 03 | 10.3 |  |
| TOTAL | 21 | 100 | 17 | 100 | 43 | 100 | 29 | 100 |  |
| alcohol Consumption |  |  |  |  |  |  |  |  | p<sup>(1)</sup> = 0.286 |
| Yes (n=44) | 7 | 63.6 | 3 | 25.0 | 17 | 48.6 | 17 | 51.5 |  |
| No (n=47) | 4 | 36.4 | 9 | 75.0 | 18 | 51.4 | 16 | 48.5 |  |
| TOTAL | 11 | 100 | 12 | 100 | 35 | 100 | 33 | 100 |  |
| Smoking |  |  |  |  |  |  |  |  | p<sup>(1)</sup> = 0.649 |
| Yes (n=49) | 9 | 60 | 6 | 42.9 | 19 | 50 | 15 | 41.7 |  |
| No (n=54) | 6 | 40 | 8 | 57.1 | 19 | 50 | 21 | 58.3 |  |
| TOTAL | 15 | 100 | 14 | 100 | 38 | 100 | 36 | 100 |  |

(\*) Significant difference at the level of 5.0%. (1) Using Pearson's Chi-square test. (2) Through Fisher's exact test. In this table, cases without information on each variable were not considered.

**Table 2:** Results of PSA tests by groups, clinical and histopathological exam.

| Variables<br>Exams | Total<br>(n= 150) |  | Atrophy<br>(n= 25) |  | PIN<br>(n= 25) |  | Quantity per group<br>BPH<br>(n= 50) |  | PA<br>(n= 50) |  | p-Value |
| --- | --- | --- | --- | --- | --- | --- | --- | --- | --- | --- | --- |
|  | n | % | n | % | n | % | n | % | N | % |  |
| **PSA** |  |  |  |  |  |  |  |  |  |  | p<sup>(1)</sup> = 0.002* |
| < 4 | 37 | 24.7 | 06 | 26.1 | 06 | 24 | 19 | 47.5 | 06 | 12.5 |  |
| 4 to 10 | 72 | 48 | 16 | 69.6 | 14 | 56 | 16 | 40 | 26 | 54.2 |  |
| > 10 | 27 | 18 | 01 | 4.3 | 05 | 20 | 05 | 12.5 | 16 | 33.3 |  |
| Uninformed | 14 | 9.3 |  |  |  |  |  |  |  |  |  |
| **Clinical exam** |  |  |  |  |  |  |  |  |  |  |  |
| Presence of nodule | 36 | 24 |  |  |  |  |  |  |  |  |  |
| Absence of nodule | 71 | 47.3 |  |  |  |  |  |  |  |  |  |
| Uninformed | 43 | 28.7 |  |  |  |  |  |  |  |  |  |
| **Histopathological** |  |  |  |  |  |  |  |  |  |  |  |
| Atrophy | 26 | 17.3 |  |  |  |  |  |  |  |  |  |
| NIP | 10 | 6.7 |  |  |  |  |  |  |  |  |  |
| HPB | 29 | 19.3 |  |  |  |  |  |  |  |  |  |
| ACP | 40 | 26.7 |  |  |  |  |  |  |  |  |  |
| ASAP | 02 | 1.3 |  |  |  |  |  |  |  |  |  |
| Others | 08 | 5.3 |  |  |  |  |  |  |  |  |  |
| Uninformed | 35 | 23.3 |  |  |  |  |  |  |  |  |  |

(\*) Significant difference at the level of 0.05 (1) Using Pearson's Chi-square test.

**Table 3:** Total PSA, free PSA, and free PSA/total PSA for each group.

| Variables | Average ± DP<br>Median (P25; P75) | | | | | p Value |
| --- | --- | --- | --- | --- | --- | --- |
|  | Total (n=150) | Atrophy (n=25) | PIN (n=25) | BPH (n=50) | PA (n=50) |  |
| Total PSA (n= 136) |  |  |  |  |  |  |
| | 10.68 ± 15.37<br>5.98<br>(3.85; 9.13) | 5.95 ± 3.12<br>5.70<br>(3.90; 7.05) | 8.22 ± 8.66<br>5.62<br>(3.70; 9.62) | 8.81 ± 16.79<br>4.41<br>(2.13; 7.26) | 15.80 ± 18.90<br>7.93<br>(5.46; 18.34) | p<sup>(1)</sup> = 0.001 * |
| Free PSA (n= 102) |  |  |  |  |  |  |
| | 1.19 ± 1.13<br>0.86<br>(0.50; 1.42) | 1.05 ± 0.37<br>0.88<br>(0.85; 1.43) | 1.21 ± 1.36<br>0.81<br>(0.40; 1.56) | 1.11 ± 1.09<br>0.82<br>(0.30; 1.37) | 1.30 ± 1.25<br>0.86<br>(0.51; 1.74) | p<sup>(1)</sup> = 0.628 |
| Free PSA/total PSA<br>(n= 102) |  |  |  |  |  |  |
| | 17.56 ± 10.29<br>16.33<br>(10.30; 23.57) | 21.20 ± 6.31<br>22.03<br>(16.30; 25.29) | 19.29 ± 14.64<br>14.93<br>(8.27; 28.64) | 19.93 ± 9.71<br>20.17<br>(15.25; 23.54) | 12.85 ± 7.53<br>11.02<br>(8.43; 17.00) | p<sup>(1)</sup> = 0.001 * |

(\*) Significant difference at the level of 0.05 (1). Kruskal Wallis test.

### Analysis of MMP-26 expression by immunohistochemistry

MMP-26 expression measured by IHC was positive in 70 (46.7%) out of the 150 samples, being more prevalent in the PA group (46/50 cases, 92%), followed by PIN (22/25 cases, 88%). The BHP group showed only 2/50 (4%) positive cases, and the atrophy group showed no reactivity (Figure 1). Weak immunostaining was observed in 34 (48.6%) of the 70 positive samples, 22 in the PIN group, 2 in the BHP, and 10 in the PC group. Moderate and intense reactivity occurred, in 31 (44.2%) and 5 (7.14%) of the positive cases, all of them in the PA group. The marking pattern was nuclear and cytoplasmic (Table 4). ROC curve analysis showed that areas under the ROC curves were larger in PA vs atrophy+PIN+BHP (AUC=0.94; 95% CI 0.9-0.98), PA+PIN vs atrophy+BHP (AUC=0.97; 95% CI 0.94-0.99) and PA vs atrophy+BHP (AUC=0.97; 95% CI 0.95-1.00) groups (Table 6; Figures 2, 3 and 4).

**Table 4.**
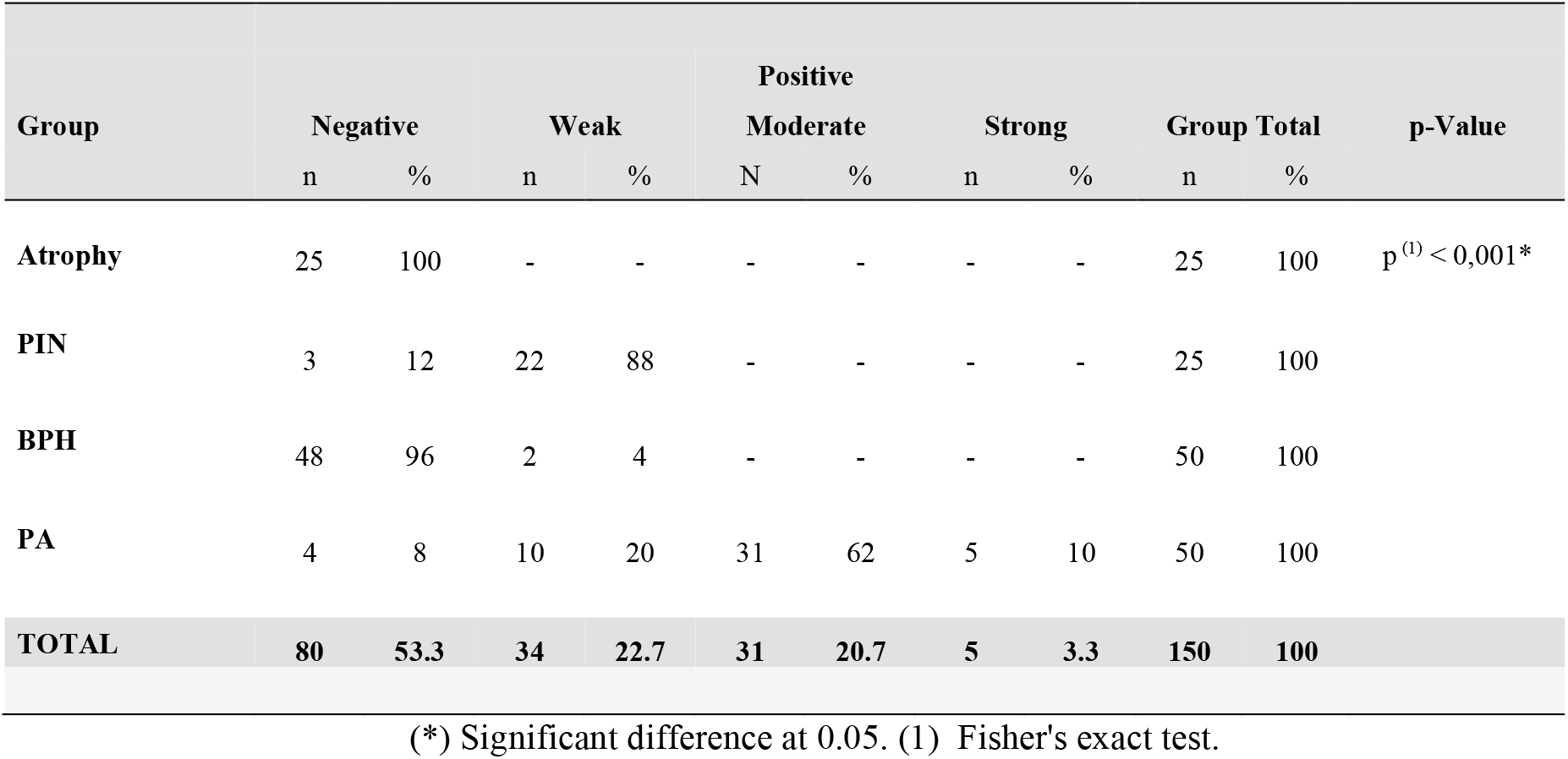
Assessment of IHC intensity for MMP26

**Figure 1.**
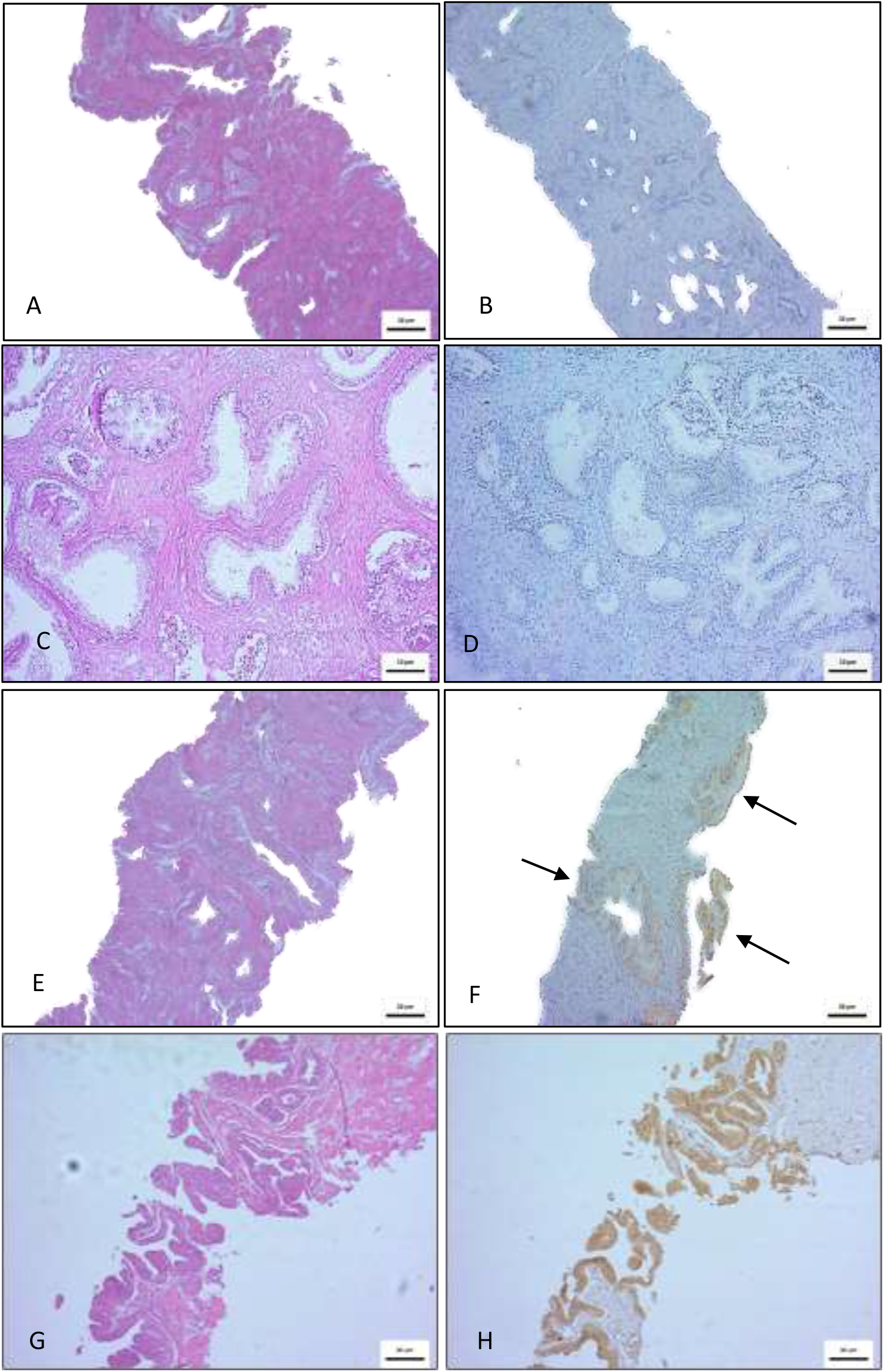
Photomicrographs of the prostate (10µM). Histopathological analysis (HE) on the left and immunohistochemistry (MMP-26) on the right. Absence of reactivity in the atrophy (A and B) and hyperplasia (C and D) groups. Weak reactivity in most cases of intraepithelial neoplasia (E and F) and moderate and intense immunoreactivity in the adenocarcinoma group (G and H). Scale bar 10 um.

**Figure 2.**
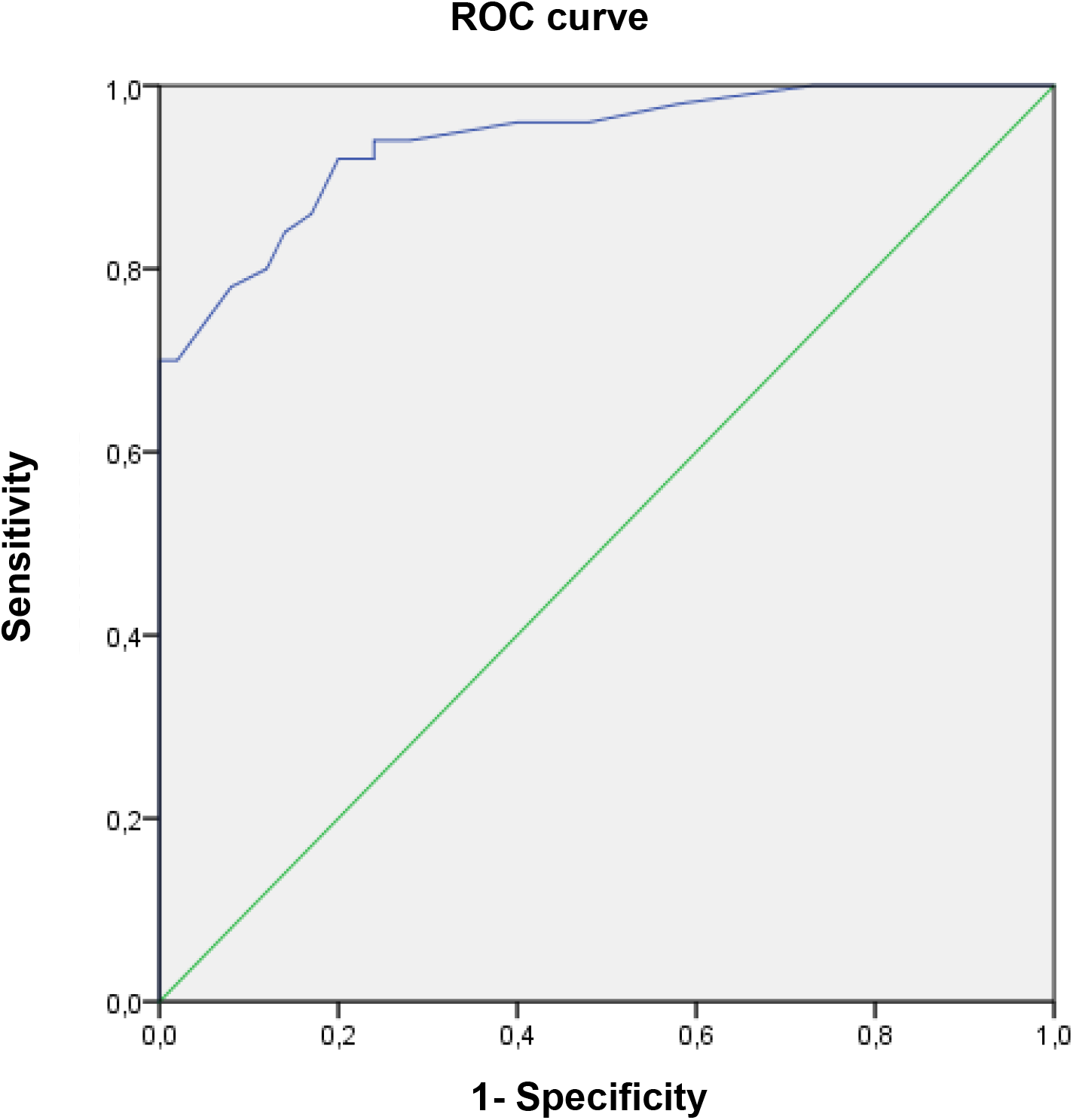
ROC curve for PA vs Atrofia+PIN+BHP regarding to the MMP-26 immunohistochemistry

**Figure 3.**
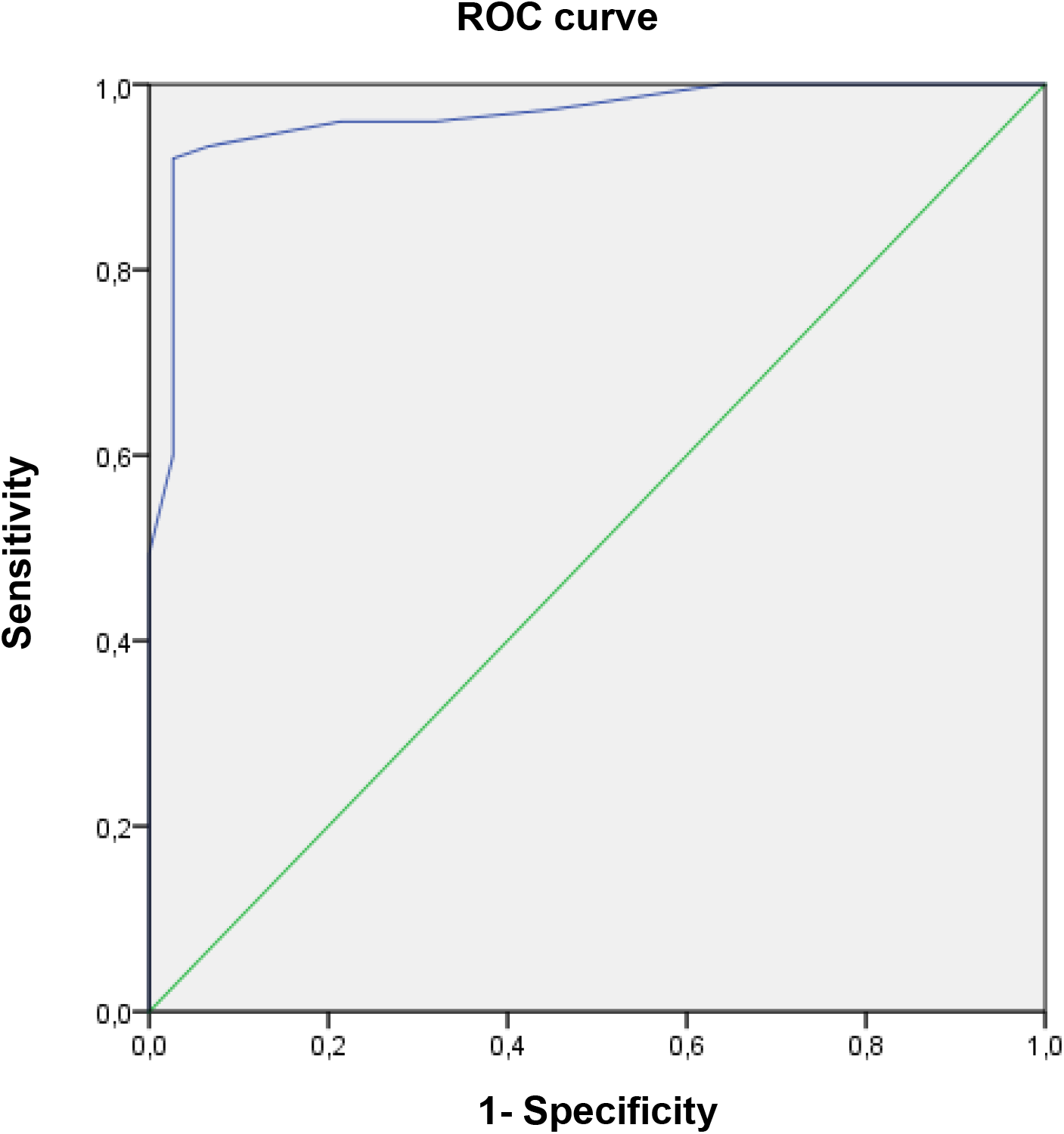
ROC curve for PA+ 521 PIN vs Atrophy+HPB regarding to the MMP-26 immunohistochemistry.

**Figure 4.**
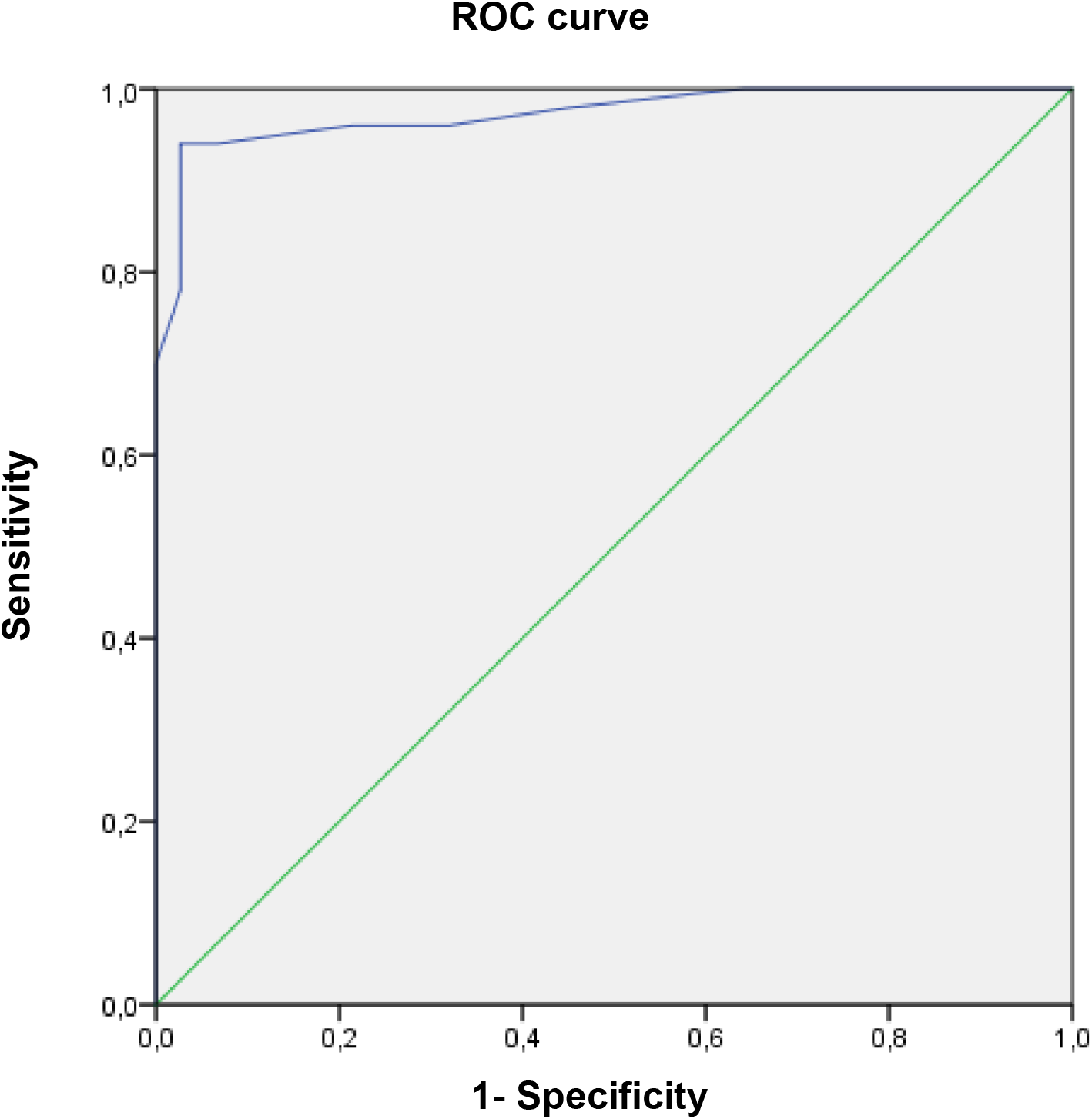
ROC curve for PA vs PIN+BHP regarding to the MMP-26 immunohistochemistry.

### Analysis of the relationship between IHC and PSA

The expression of MMP-26 by IHC showed a significant association with total PSA values. The greatest difference occurred in the PSA group with up to 4 ng/ml, where most cases showed negative IHC (38.6%), followed by the PSA group greater than 10 ng/ml, where values were higher among those who had positive IHC (28, 8%). An association was also found between the intensity of the reaction and PSA values. There was a variation of 15% to 17% between the weak and moderate intensities in the PSA group with up to 4 ng/ml and 40% in the intense reaction for the PSA group greater than 10 ng/ml (Table 5).

**Table 5:**
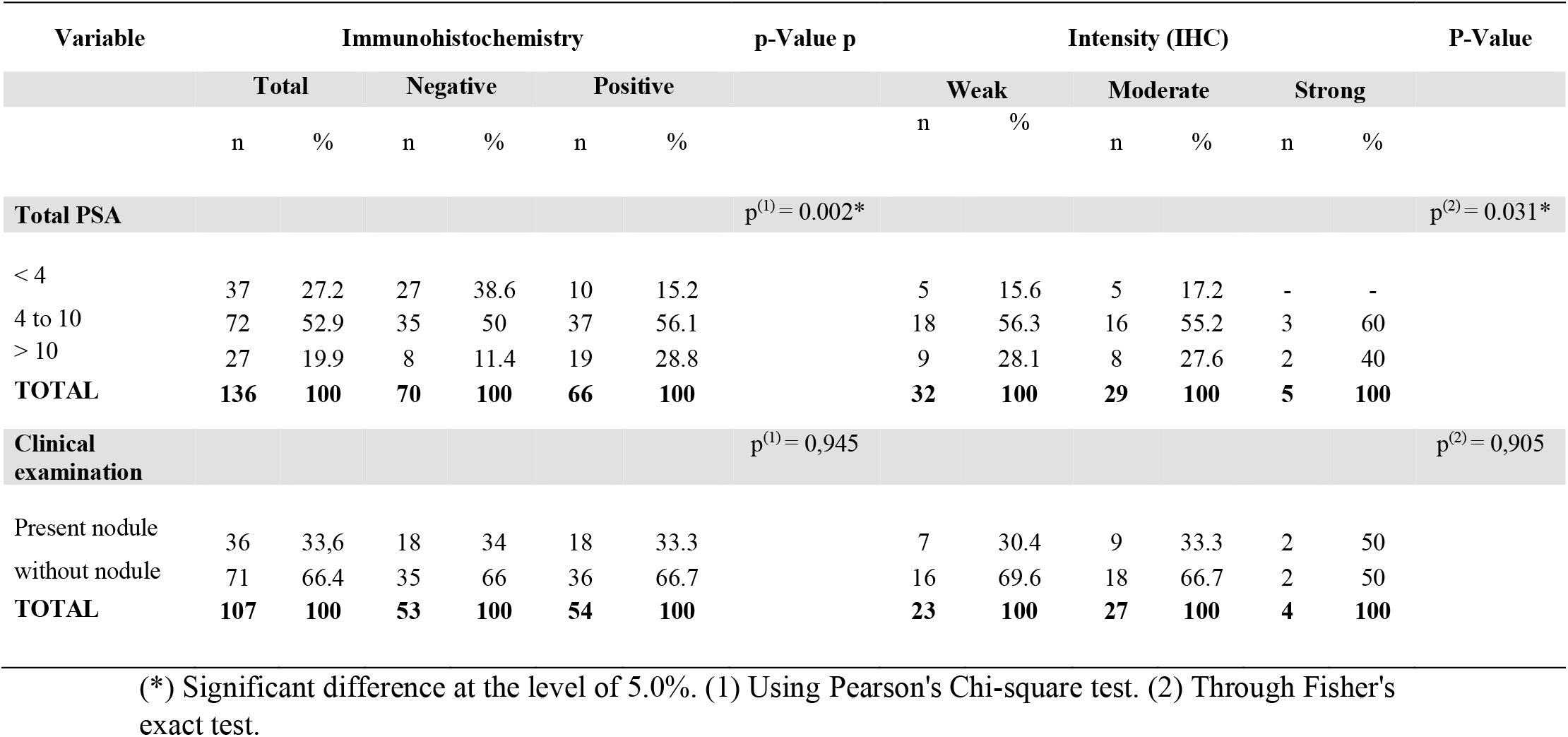
Evaluation of PSA and clinical exams according to IHC

**Table 6.**
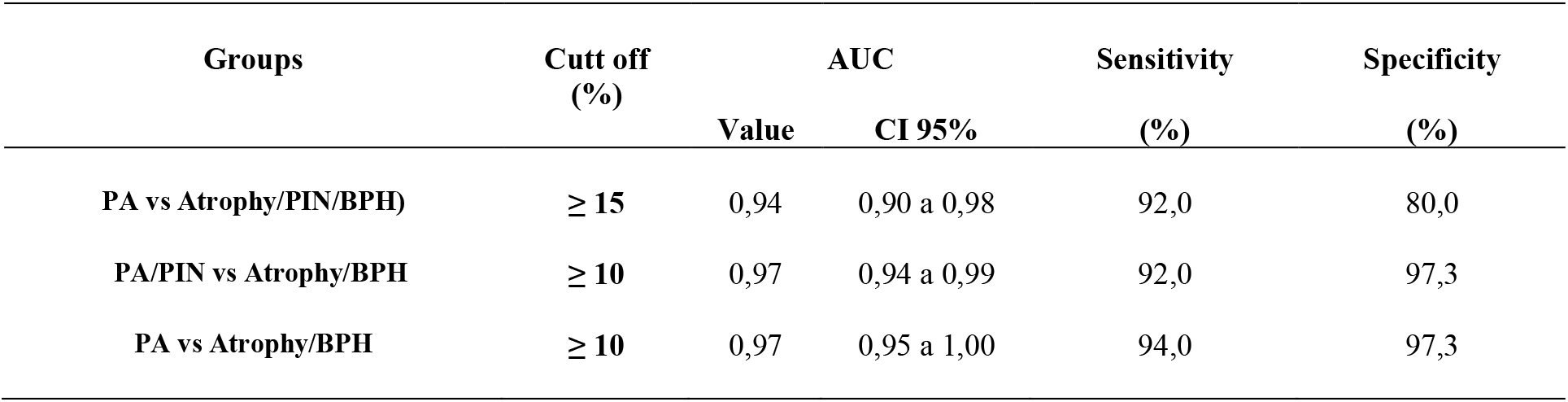
ROC analysis of groups regarding to the MMP-26 IHC

## DISCUSSION

Since the 1980s, with the introduction of the PSA test in the medical routine, there has been a significant increase in the diagnostic of PC cases and a 20% reduction in the PC mortality rate. Serum PSA is measured for early detection, staging, and monitoring of the disease. However, it might have low specificity when the values range between 3 and 10 ng/ml, and also showing an increase in benign conditions such as BHP and prostatitis ^2,16^. The search for new biomarkers, which combined with PSA can provide greater precision in the diagnosis, has led to several studies on the expression of MMP, both in serum and in tissue samples of PC.

Kanoh et al. (2002) and Zhang et al. (2004) observed, high serum levels of MMP-2 and MMP-9 in patients with PC and BHP, with increasing expression as the disease progressed ^17,18^. In addition to MMP-2 and -9, Morgia et al. (2005) found that MMP-13 levels were significantly higher levels in patients with PC ^19^. MMP-7 was analyzed by Szarvas et al. (2011), and its levels were also strongly elevated, especially in patients with metastatic PC ^20^. In 2005, Riddick et al. evaluated the expression of MMP-10, -15, -24, - 25, and -26, of which MMP-26 was the one showing the highest degree of expression in malignant prostate tissues, strongly correlating with the Gleason score ^21^.

Recently, Cheng et al. (2017) observed a marked increase and strong immunoreactivity of MMP-26 in the serum, in PC samples compared to the BHP and control groups ^16^. Also, Zhao et al. (2003) noted that MMP-26 was able to activate MMP-9 in PC cells, an essential mechanism in the invasion and metastasis process ^22^. An assay with specific inhibitors for MMP-26 generated a reduction in MMP-9 and a significant decrease in the invasive potential of these cells. These findings support the hypothesis that the activation of MMP-9 by MMP-26 may promote an increase in cell invasiveness and, consequently, a worse clinical outcome.

Our results also demonstrated a higher expression of MMP-26 in cases of PA (63%), in addition to a positive association between the IHC expression of MMP-26 with PSA values, which suggests that this enzyme could be used as an adjunct to the measurement of this marker for the differential diagnosis between PA and BHP, as well as for estimating the prognosis of cancer given that it has also shown weak reactivity in NIP samples. The rupture of continuity between the cell layer and the basement membrane is essential for the progression from intraepithelial neoplasia to prostate carcinoma. Lee et al. (2006) found a higher expression of MMP-26 in cases of PIP compared to PC and that its levels declined in the progression to invasive cancer ^23^. His findings suggest that MMP-26 might play an important during tissue transformation between these lesions, serving as an early marker of prostate cancer.

The prognosis of PC is also associated with age, with older age being the best-established risk factor. Studies indicate that both incidence and mortality increase after age 50, with three-quarters of new cases around age 65 ^2,16^. In this context, we observed that 56% of the patients with PA in our study were between 60 and 69 years old. Moreover, our findings showed total negativity for MMP-26 in the group of atrophic cases, which might be useful for therapeutic definition as a predictive marker in such patients who also have high PSA. Furthermore, MMP-26 can be measured in serum, as described by Cheng et al. (2017) in their study ^16^. Thus, it would be possible to concomitantly evaluate PSA and MMP-26 in the same blood sample and estimate the outcome of patients at risk in a given age group.

Ethnicity can also be a risk factor, blacks are more susceptible, according to a study by Noone et al. (2017), which contradicts our results: 44% of the samples were obtained from mixed races and 19% from blacks (with 10% presenting CP) ^24^. It is known that smoking promotes an increase in mortality among patients with CP ^25^. However, this association was also not found and can be explained by the case selection being based on histopathological alteration and not spontaneous demand.

Unlike most MMPs, MMP-26 is expressed in the intracellular region ^8^. The cytoplasmic immunoreactivity of MMP-26 was evidenced by Guo et al. (2018) in glioma cells and by Hu et al. (2014) in colorectal carcinoma ^15,26^. We observed both a cytoplasmic and nuclear pattern in malignant PC. This differentiated location enables new roles for this enzyme. Although the expression of MMP-26 is elevated in several types of cancer such as liver, lung, and breast cancer ^9–11^, some studies have also revealed protective functions in the regulation of inflammation and apoptosis ^8,27,28^.

Khamis et al. (2013) suggested an anti-inflammatory role for MMP-26 ^8^. Cells transfected with MMP-26 cDNA showed low regulation of inflammatory genes, whereas cells with the silenced gene had reduced IL-10 receptor (IL10R), suggesting that MMP-26 deficiency may promote inflammation by inhibiting the IL10R pathway. Subsequently, Khamis et al. (2016) provided evidence for a protective role in the prostate ^27^. It was observed that prostate tumor cells expressing MMP-26 underwent apoptosis via Bax and that MMP-26 overload would have a pro-apoptotic function, reducing the degree of invasion and disease progression. It was also observed by Savinov et al. (2006) that the elevation of MMP-26 resulted in a favorable prognosis and increased survival of patients with breast carcinoma *in situ* ^28^.

Given the versatile profile of MMP-26, there is still much to be investigated. However, several studies have demonstrated its potential as a tumor marker, especially for prostatic carcinoma. In this study, we showed that the MMP-26 IHC can be potentially useful to differentiate benign and malignant cases of the prostate, and may assist in the development of new strategies for the diagnosis, prognosis, and treatment of prostate cancer.

## Data Availability

The authors confirm that the data supporting the findings of this study are available within the article.

## ACKNOWLEDGMENTS

This work was supported by CAPES (Coordination for the Improvement of Higher Education Personnel), LIKA (Keizo Asami Immunopathology Laboratory), and LPCM (Cytological and Molecular Research Laboratory).

## AUTHORS ’CONTRIBUTION

All authors contributed to the conception and design of the study. The collection and processing of the material, as well as the immunohistochemical analyzes, were performed by Romildo Luciano da Silva, Francisco Luís Almeida Paes, and Sandra Maria Souza da Silva. The analysis of the data and results obtained was carried out by Jacinto Costa da Silva Neto. Ingrid Tavares de Lima did the drafting of the manuscript. The revision and editing of the manuscript were carried out by Ana Pavla Almeida Diniz Gurgel and Jacinto Costa da Silva Neto. The financing was acquired by Jacinto Costa da Silva Neto. The research was supervised by Jacinto Costa da Silva Neto. All authors read and approved the final manuscript.

## APPROVAL OF ETHICS

The Research Ethics Committee approved this study of the Health Sciences Center of the Federal University of Pernambuco (CEP / CCS / UFPE) under CAAE: 79701517100005208

